# Unlocking uterine biology at home: a validated system for DNA, RNA, and microbial analysis from menstrual effluence

**DOI:** 10.1101/2025.05.21.25327630

**Authors:** Stephen Gire, Xitong Li, Claire Toth, Meet Doshi, Sachin Kumar Gupta, Scott Parker, Debbie Boles, Ridhi Tariyal

## Abstract

**Introduction:** Access to accurate, non-invasive diagnostics remains a critical unmet need in women’s health. Menstrual effluence, containing endometrial tissue, immune cells, and microbial communities, represents a clinically relevant specimen for genomic and molecular pathology applications, yet has historically been underutilized due to concerns about sample integrity and variability.

**Methods:** We developed and validated a standardized, at-home tampon-based collection system designed to preserve nucleic acids at ambient temperature for clinical-grade analyses. 1,067 tampon samples from 328 participants underwent, RNA sequencing and metatranscriptomic profiling to assess specimen transcript integrity, diagnostic fidelity, and microbial composition over time. 12 patients were exome sequenced using matched menstrual effluence and whole blood to assess assay concordance between sample types.

**Results:** RNA extracted from menstrual effluence maintained stability for up to 14 days without refrigeration, achieving sufficient yield and quality for sequencing in >97% of samples. Variant detection via exome sequencing demonstrated 100% concordance among overlapping single nucleotide variants between menstrual fluid and matched venous blood, confirming clinical equivalency for genetic testing. Transcriptomic analyses revealed cycle-dependent variation in key reproductive and immune markers, while metatranscriptomic profiling identified shifts in microbial communities consistent with known reproductive tract dysbiosis.

**Conclusions:** Standardized at-home collection of menstrual effluence provides a clinically actionable platform that supports remote specimen acquisition without compromising molecular assay fidelity, offering a scalable solution to improve access to carrier screening, reproductive health assessment, and infectious disease monitoring in clinical practice.

## INTRODUCTION

Women’s health has been underserved by non-specific diagnostics and limited therapeutic options. Despite high disease burden, conditions like endometriosis, fibroids, and recurrent miscarriage are diagnosed and treated with tools that lack precision, leading to significant delays in care^1^. Broader reliance on non-specific clinical pathways has led to a one-size-fits-all approach to care^2–4^. For instance, gonadotropin-releasing hormone (GnRH) analogs—despite inducing premature menopause and causing severe side effects such as bone loss and mood disturbances^5^ – remain a first-line treatment for endometriosis^6^ and fibroid-related heavy bleeding^7^. CA125, a non-specific marker of inflammation, is monitored across a wide range of conditions including ovarian cancer^8^, heart failure^9^, miscarriage risk^10^, and endometriosis^11^, despite limited diagnostic specificity^12^. Without condition-specific diagnostics, targeted therapies have remained underdeveloped, perpetuating inadequate care across puberty, pregnancy, and menopause.

At-home biospecimen collection systems offer a scalable way to improve diagnostics and enable precision medicine in women’s health. Remote collection of biological samples allows clinical data to be captured outside of brick-and-mortar healthcare settings, improving access and enabling earlier decision-making^13^. For research, these systems offer opportunities to build rich, longitudinal, high-dimensional datasets that are essential for studying reproductive biology at scale. However, few such datasets exist, and those available are poorly harmonized^14^, limiting their cross-study comparability and value for biomarker discovery.

Menstrual effluence is a rich but underutilized biospecimen that closely mirrors the endometrial environment and presents novel opportunities for diagnostic development. Menstrual fluid is expelled naturally and contains a mix of shed endometrium, interstitial fluid, blood, mucosa, and epithelial cells from the cervix and vagina, along with reproductive tract microbiota^15,16^. Organoids derived from menstrual effluence demonstrate similar transcriptomic profiles and hormone responsiveness as those from invasive endometrial biopsies^17^. Importantly, menstrual effluence contains a unique population of immune cells enriched for innate and adaptive subsets involved in tissue remodeling and immune tolerance^18^, unlike peripheral blood. The sample’s heterogeneity captures a broader snapshot of uterine and vaginal health than traditional biopsies or swabs.

Despite its promise, menstrual effluence has historically been viewed as technically difficult to work with. Approximately 30% of cells in menstrual fluid are non-viable^15,16^, and tissue aggregates and mucosal debris complicate laboratory workflows. Additionally, cellular lysis releases nucleases that can degrade nucleic acids, compromising downstream molecular assays. Sample composition also varies by collection method^19^ – e.g., pad vs. tampon vs. menstrual cup – introducing variability and contributing to skepticism about its reliability as a biospecimen. However, these challenges are surmountable with proper pre-analytical protocols.

New advances have transformed how menstrual effluence is viewed in diagnostics and research. Recent studies have generated high-resolution datasets from menstrual fluid and uterine tissue that offer insight into reproductive pathophysiology^15,16,20^. Transcriptomic data from menstrual effluence have revealed dynamic biological processes such as epithelial-to-mesenchymal transition relevant to endometriosis progression^21^. Endotoxins detected in menstrual samples have been linked to adverse pregnancy outcomes^22^, and menstrual immune profiles have shown potential for diagnosing inflammatory and infectious conditions^23^. These discoveries demonstrate that menstrual effluence is not only clinically relevant but may offer information inaccessible via other sample types.

We developed a standardized, tampon-based system to enable high-quality menstrual fluid collection at home. To address pre-analytical variability and improve sample stability, we built a self-contained tampon collection kit that also captures metadata such as cycle day and bleeding volume, which serve as important co-variables in transcriptomic analyses. Prior research has shown that aligning sample timing with menstrual phase improves diagnostic accuracy in cardiovascular^24^ and mental health applications^25^. The NextGen Jane® (NGJ) platform enables consistent sample collection, better supports hormonal context interpretation, and allows for widespread use in both clinical and research settings.

We validated our tampon-based sample collection system through large-scale deployment and demonstrated its feasibility and diagnostic utility in three clinically relevant applications through DNA exome sequencing (carrier screening) and unbiased RNA sequencing of human and microbial cell types (molecular phenotyping and microbial surveillance). We have collected 2,025 RNA-seq and metatranscriptomic datasets from 1,067 tampons submitted by 328 U.S. participants. All samples were home-collected and shipped at ambient temperature to a centralized lab in Oakland, CA, without loss of data quality – demonstrating the viability of decentralized sampling for molecular diagnostics.

## MATERIALS AND METHODS

A thorough accounting of materials and methods can be found in Supplemental Methods.docx

### Patient Recruitment and Metadata Collection

Patients were recruited under IRB-approved protocols (20192619, 20191947, 20233438). Consent was obtained from participants who agreed to tampon-based specimen collection and completed a survey on clinical history, menstrual regimen, and demographics. Data were organized in four tiers: patient-level (race, ethnicity), cycle-level (age, geography, birth control), tampon/kit-level (flow type, cycle day), and sample-level (RNA concentration, library size). Sample accession and collection dates were recorded to assess sample stability over time. Flow rate was calculated from the weight of the tampon before and after collection. Metadata can be found in Supplemental_RNASeq.xlsx. Patient IDs C01-, H01, E01), Kit IDs (NAPS) and Sample IDs (ESGs), are randomized and anonymized ID’s that cannot be tied back to patients outside the research team.

### Sample Collection and Processing

Participants used organic cotton tampons during multiple days of menstruation, wearing them for 4 hours. After collection, the tampon was sealed in a collection jar with 20mL of preservation buffer (Norgen Biotek). Samples were sent back to the lab via USPS. Upon receipt, samples were processed, including extrusion, centrifugation, and aliquoting into cryovials for storage at −80°C.

### Hemoglobin Content Spectrophotometry

The tampon sample was diluted 1:4 with a preservation buffer and analyzed using a UV/Vis spectrophotometer to measure absorbance at 550nm.

### Nucleic Acid Extraction and Quality Assessment

RNA and DNA were extracted using the Norgen Column-Based RNA Extraction and MagMax mirVana Total RNA Isolation methods. For RNA, DNase treatment was applied, followed by clean-up with RNA XP clean beads. DNA was extracted using the MagMax-96 DNA Multi-Sample Kit. The extracted nucleic acids were quantified using a Qubit 4.0 fluorometer.

### Exome Sequencing and Carrier Screening

DNA exome sequencing was outsourced to Labcorp for analysis under their established protocols. SNVs and sample metadata can be found in Supplemental_DNASeq.xlsx

### RNA Sequencing and Analysis

RNA libraries were prepared using the Zymo-Seq RiboFree Total RNA Library Kit, followed by sequencing on an Illumina NextSeq2000. Data processing included adapter trimming, alignment to the hg38 human genome, and gene count normalization using STAR Aligner and FeatureCount. RNA degradation effects were accounted for using the DegNorm program. Sequencing metadata and gene expression values can be found in Supplemental_RNASeq.xlsx. Tampon and menstrual cup samples were compared to publicly available RNA sequencing data on GTeX to calculate Spearman correlations between tissue types. Samples used for this analysis can be found in Supplemental_GTeX.xlsx. Differential expression was performed in XLSTAT.

### Microbiome Analysis

Metatranscriptomic reads were aligned to microbial databases using Kraken2 to generate raw read counts. For microbiome diversity, species-level and genus-level counts were aggregated to genus level and analyzed using log-transformation and CPM normalization. Sequencing metadata and gene expression values can be found in Supplemental_RNASeq.xlsx

### Metadata and Statistical Analysis

Metadata, including clinical phenotypes and sample characteristics, were used in ANOVA to assess statistical significance across various groups. Both pvalue and cohen’s f for effect size were calculated for all analyses using XLSTAT. Analysis focused on bleeding phenotypes, flow rates, and other collection-related variables.

## RESULTS

### A tampon-based at-home collection system can reproducibly collect menstrual effluence for downstream molecular analyses under real-world mailing and storage conditions

The NGJ tampon collection system was designed for ease-of-use, integration into daily routines, and standardization to ensure reliable biological preservation. Participants use the provided TOTM-brand organic, low-absorbency tampons, minimizing synthetic additives and heavy metals. Tampons are worn for four hours – reducing toxic shock risk and facilitates sample processing. The cardboard applicator reduces contact with the absorbent core, limiting commensal contamination. A nitrile glove is included for removal and placement into the jar. Once sealed, the cap pulls the string into the jar, preventing leaks, and a shuttle punctures a foil seal to release the preservation buffer, supersaturating the tampon. The sample is bagged, intake questions completed and returned via standard mail. User-tested instructions guide participants through the collection process. A picture of the collection kit and cover of the instructions for use can be found in Figure 1.

**Figure 1:**
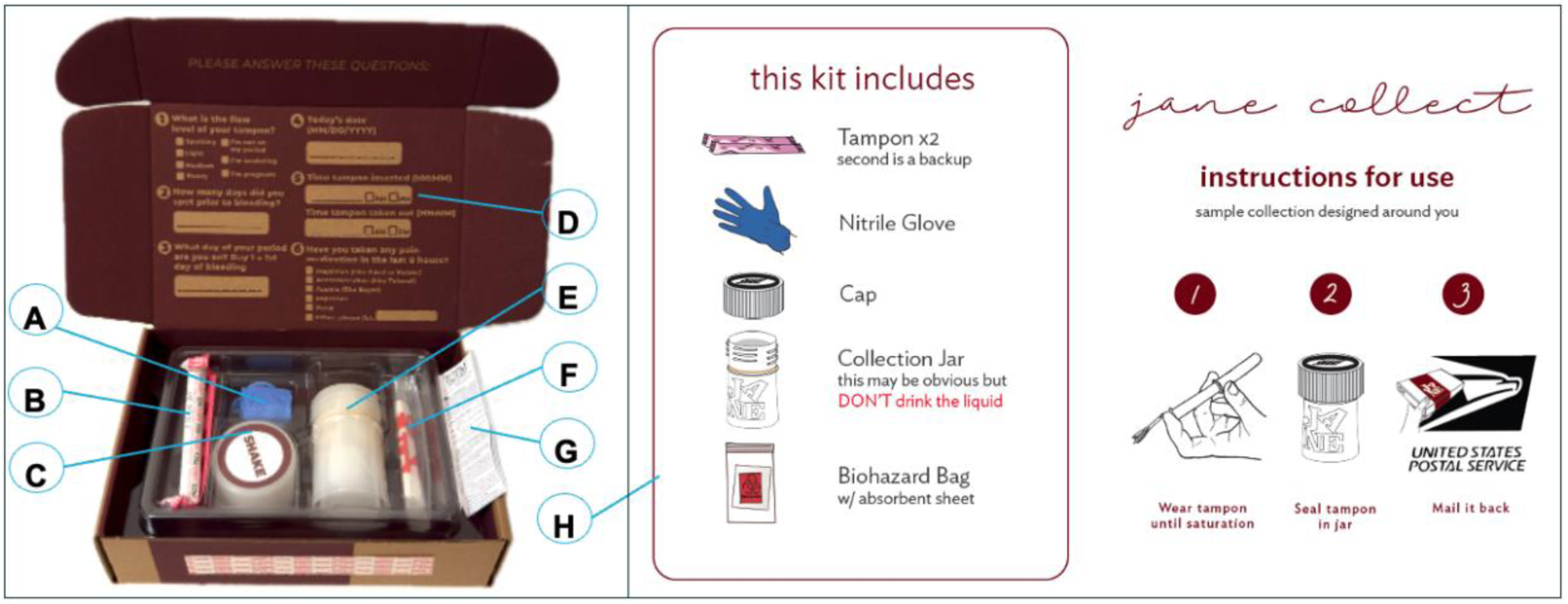
NextGen Jane tampon collection kit. NextGen Jane collection kit. **a.** Nitrile glove included for tampon removal. **b.** Included tampon with applicator. **c.** Cap that seals tampon in jar and releases preservative onto tampon. **d.** Basic phenotypic data about current collection is recorded on the inside flap of the box. These include the flow type of the tampon being collected, the day of the cycle, the date, time in and time out for collection, as well as a list of any medications taken during the collection **e.** Collection jar includes a sealed buffer cup with preservative. **f.** Biohazard bag with absorbent material for shipping. **g.** Tampon instructions for use. **h.** Front cover of the tampon collection instructions for use.

*Demographic Data of Sample Population –* We analyzed menstrual samples collected on cycle days 1–3 with complete phenotypic and technical data (Supplementary Table 1). Inclusion criteria ensured samples were menstrual (not off-cycle vaginal tampons), had complete metadata (flow type, collection time, bleeding phenotype), and passed sequencing quality checks (strandedness, ribosomal rRNA content and uniquely mapped reads). This resulted in 1,135 sequences from 601 tampons across 276 participants.

Table 1 summarizes participant demographics and key flow characteristics. 75% (207 participants) identified as Non-Hispanic White, with the remaining 69 participants identifying as Asian, Black, or more than one race. Of 365 cycles, 287 were from individuals not using birth control. Enrolled participants were asked to report the ***flow phenotype*** (overall period heaviness), characterized as heavy, medium or light, corresponding to their subjective experience of their entire menstrual cycle at the time of collection. ***Flow type*** (heaviness on the collection day) was also reported by participants. These measures differ – e.g., a heavy-flow tampon from a light bleeder may differ significantly from one of a heavy bleeder.

**Table 1:** Patient, Cycle, and Tampon level data collection summary. Data was collected at the **a.** patient level, as well as the **b.** cycle and **c.** tampon level. Each patient may have multiple collections within a cycle, and between many monthly cycles. During this time of collection, patients age, change birth control, or may change their bleeding phenotype. We have collected data on these three hierarchical levels to reflect these changes. This dataset includes 601 tampons collected in 365 monthly cycles from 276 participants.

| A. Patient Level Data for 276 Patients | B. Cycle Level Data for 365 Cycles | C. Tampon/kit level Data for 601 Tampons |
| --- | --- | --- |
| **Age** | **Age** | **Season of Collection** |
| <35 166 | <35 218 | Spring 223 |
| 35-40 74 | 35-40 99 | Summer 157 |
| >40 36 | >40 48 | Autumn 135 |
| **Ethnicity** | **Birth Control** | Winter 86 |
| Hispanic of Latino 33 | No birth control 287 | **Flow Type** |
| Non-Hispanic 235 | Progestin 28 | Heavy 294 |
| Other/NA 8 | Estrogen 2 | Medium 183 |
| **Race** | Estrogen and Progestin 41 | Light 113 |
| Asian 23 | Other 7 | Spotting 11 |
| Black or African American 21 | **Bleeding Phenotype** | **Cycle day** |
| More than on race 21 | Heavy 170 | 1 172 |
| White 207 | Medium 154 | 2 284 |
| American Indian or Alaska Native 1 | Light 41 | 3 145 |
| Other/NA 3 |  |  |
| **Region of Collection** |  |  |
| Midwest 42 |  |  |
| Northeast 46 |  |  |
| South 91 |  |  |
| West 97 |  |  |

### Menstrual effluence preserved in stabilization buffer maintains sufficient RNA integrity for sequencing for at least 14 days at ambient temperature

Home-based sample collection enables large-scale, longitudinal studies across diverse locations. We report nucleic acid yields from our tampon collection system and transcript degradation index (DI), similar to the transcript integrity number (TIN)^26,27^. For sequencing, 11% of samples failed at least once. Among initially failed samples, 79% were successfully resequenced, yielding a final estimated sample failure rate of 2.3%. Approximately 60 million reads are generated per sample (median = 60 million ± 23 million reads), which include both human and microbial transcripts.

Nucleic acid extracted from a 600ul aliquot of sample yields enough RNA for downstream sequencing. After lysate release from tampons, an average of 17 mL of total volume (mean = 17.13 ± 2.73) was recovered, providing sufficient material for downstream assays. A 500 ng RNA threshold – required for Zymo RiboFree sequencing – was routinely achieved. Although this threshold is reachable via deep-well extraction, optimizing magnetic bead protocols significantly improved nucleic acid recovery (Table 2), enabling use of standard plate formats. Using 600 µL of lysate, 500 ng RNA was obtained in 93% of samples, with a median RNA yield of 1.5 µg.

**Table 2:**
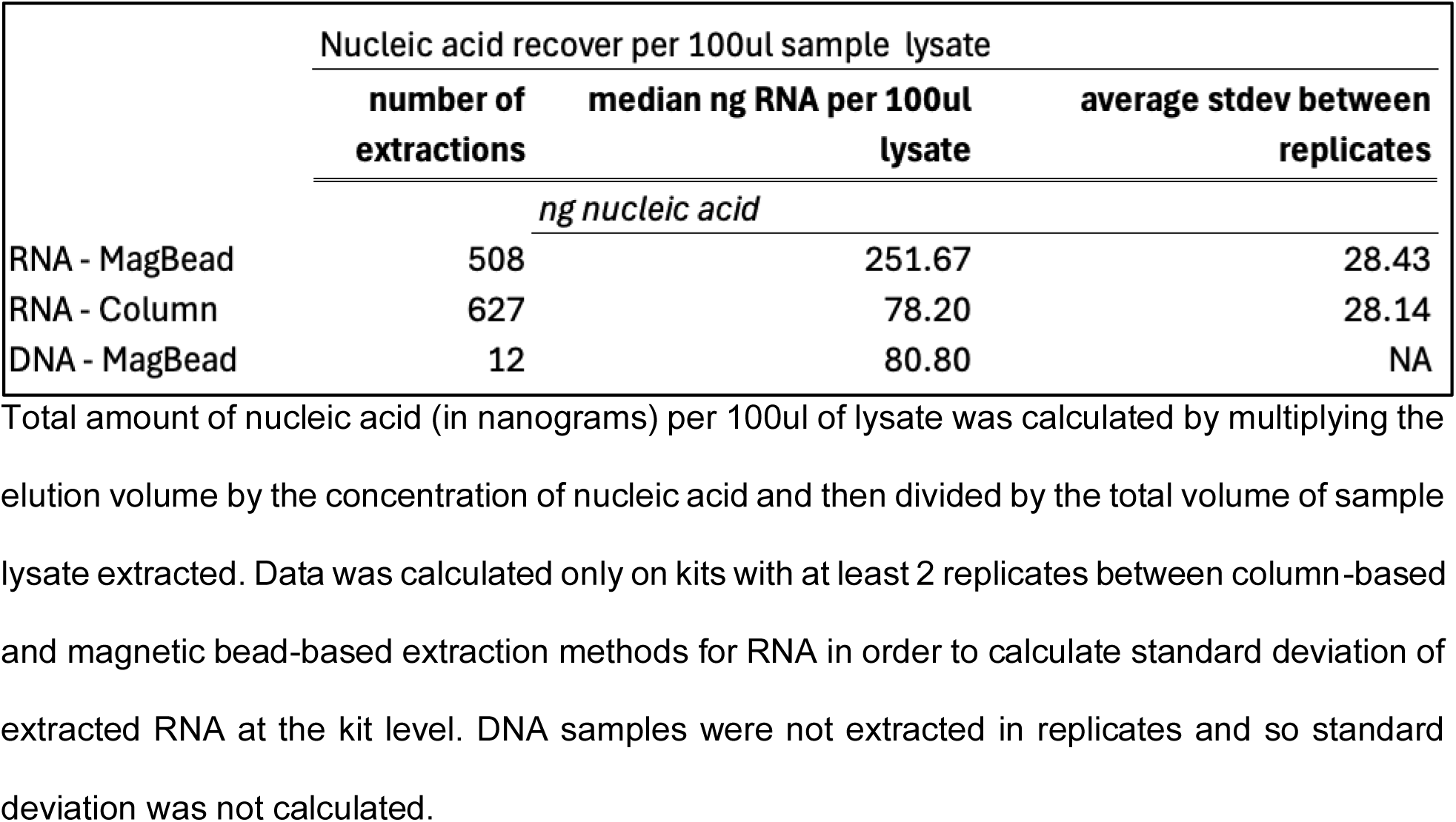
Nucleic acid recovery statistics. Total amount of nucleic acid (in nanograms) per 100ul of lysate was calculated by multiplying the elution volume by the concentration of nucleic acid and then divided by the total volume of sample lysate extracted. Data was calculated only on kits with at least 2 replicates between column-based and magnetic bead-based extraction methods for RNA in order to calculate standard deviation of extracted RNA at the kit level. DNA samples were not extracted in replicates and so standard deviation was not calculated.

We assessed transcript degradation during ambient-temperature storage by measuring the DI of the 25th percentile of genes using DegNorm^26^. RNA degradation causes 3′ coverage bias in RNA-seq data. The DI ^26^ quantifies this degradation at the gene level, with higher scores reflecting greater 3′ bias. Based on sample collection and processing dates, samples spent an average of 6.7 ± 3.52 days in buffer. No significant change in 5′–3′ coverage bias (DI_25) between days 1 and 21 (Figure 2a), indicating stability at typical shipping conditions. We further assessed the impact of menstrual flow rate on RNA quality. Flow rate had a strong effect on DI_25: participants with slower flows (<1 grams/hr) had higher degradation, whereas faster flow rates (1–2 grams/hr or >2 grams/hr) correlated with reduced degradation (p=0.002; cohen’s f = 0.230 Figure 2b). Extended collection times in slow-flow individuals may exacerbate degradation.

**Figure 2:**
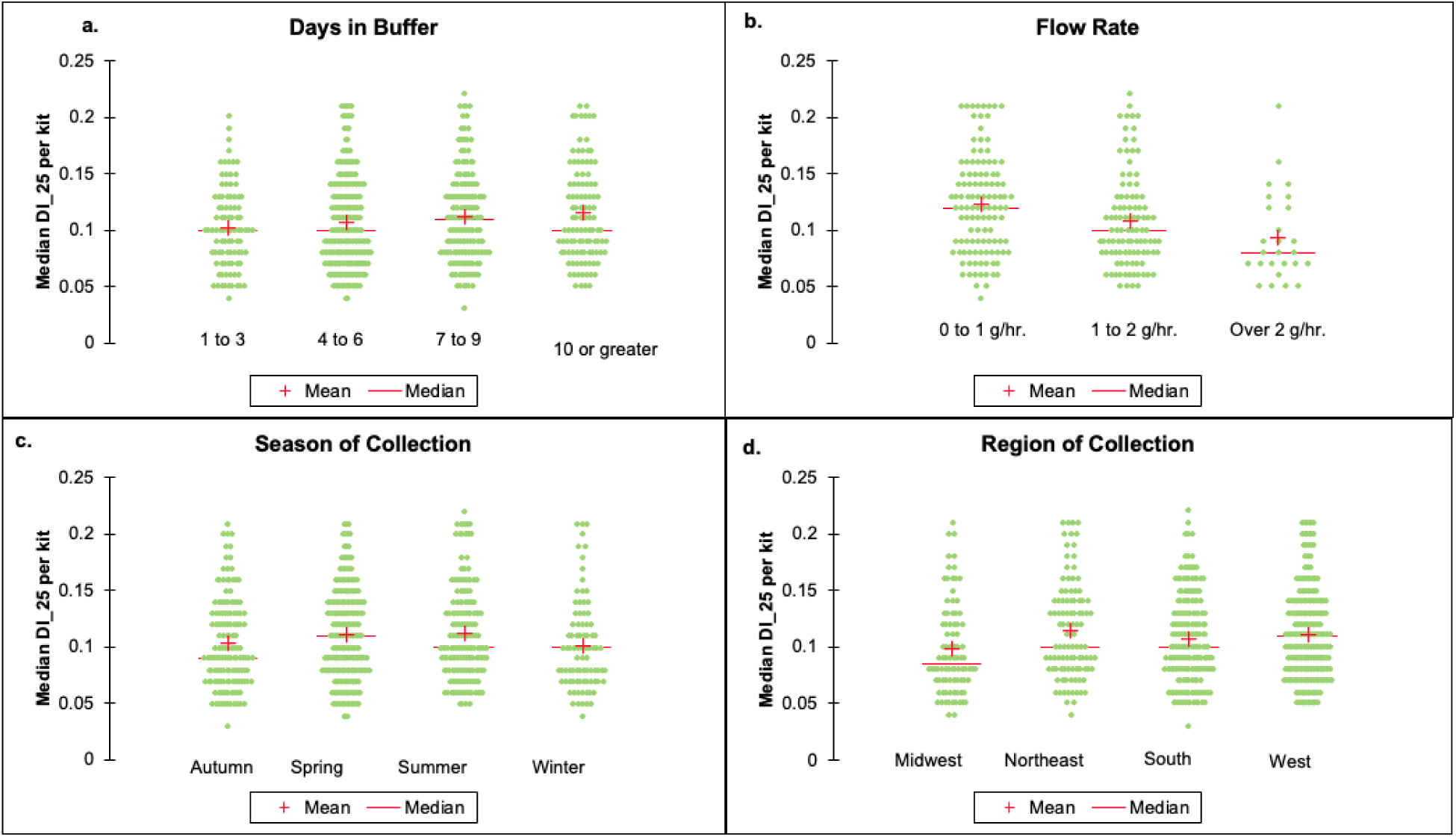
Degradation Index and sequence library size performance metrics. Sequencing metrics for monitoring degradation of RNA transcripts in menstrual collections. Higher DI scores correlate to more degraded samples. **a.** DI_25 score variance grouped by days sample left in buffer during transit show no significant changes by day (p = 0.107; cohen’s f = 0.101). Samples were grouped days 1-3, 4-6, 7-9, and 10 or greater. **b.** DI_25 score plotted by rate of menstrual flow (grams of fluid collected divided by the hours of collections. Samples are bucketed in 0 to 1 grams of fluid lost per hour, 1 to 2 grams lost per hour, and over 2 grams lost per hour. There was a significant decrease in degradation as flow rate increased (p=0.002, cohen’s f = 0.230) **c.** & **d.** DI_25 plotted by season (U.S) and by region (U.S.) the collection occurred. Midwest collected samples showed a lower mean DI_25 compared to other regions (p = 0.023; cohen’s f = 0.127). Analysis was performed at the individual sequence level (n=1,135)

No seasonal effects on DI_25 were observed (Figure 2b). We did detect significantly lower overall degradation in samples collected in the Midwest region of the United States (p = 0.023; cohen’s f = 0.127) (Figure 2d). While not statistically significant, samples from the Midwest also show an increase in mean flow rate compared to other regions (Midwest mean= 1.423 g/hr; Other Region mean = 1.149 g/hr; p = 0.090; cohen’s f = 0.110). This could explain the decrease in DI_25 scores for this region. A total of 11 kits were successfully sequenced at least three times by separate technicians, showing a median coefficient of variation between replicates of 0.149 (supplemental data).

From 600 µL lysate, we consistently recovered sufficient RNA and DNA for sequencing. RNA remains stable for at least two weeks in buffer at ambient temperatures, showing minimal transcript degradation (see Figure 2a,b) – making this system ideal for at-home collection. With 17 mL total lysate per tampon (equivalent volume to extract and sequence the sample 28 times), ample material remains for molecular assays and biobanking. Collectively, we have biobanked 2,013 tampon-derived samples.

### DNA from tampon-collected menstrual effluence yields variant calls that are concordant with those obtained from matched blood-based exome sequencing

At-home, longitudinal collection of menstrual effluence offers accessible prenatal and antenatal screening, especially in gynecological deserts or where remote sampling enhances clinical care^28^. To assess concordance of single nucleotide variants (SNVs) between menstrual effluence and venous blood, both sample types were collected from 12 participants and exome sequenced via Labcorp’s Inheritest.

Percentage of overlapping SNVs between blood and menstrual samples was high (mean = 0.983 ± 0.0089), which is similar to other studies showing high concordance of SNVs between different sample types from the same patient^29^. On average, menstrual effluence detected 143 more SNVs than venous blood. Non-overlapping SNVs had lower coverage than overlapping SNVs, likely explaining the difference (median coverage depth for overlapping SNVs = 103 vs. non-overlapping SNVs = 40). Four participants carried pathogenic variants detected in both sample types: GJB2:p.Gly12fs (hearing loss), CFTR:p.Ile507_Phe508delinsiIle (cystic fibrosis), CAPN3:p.Arg489Gln (limb-girdle muscular dystrophy), and HEXA:p.Arg499Hi (Tay-Sachs), with allele frequencies ranging from 38% to 62% (Figure 3).

**Figure 3:**
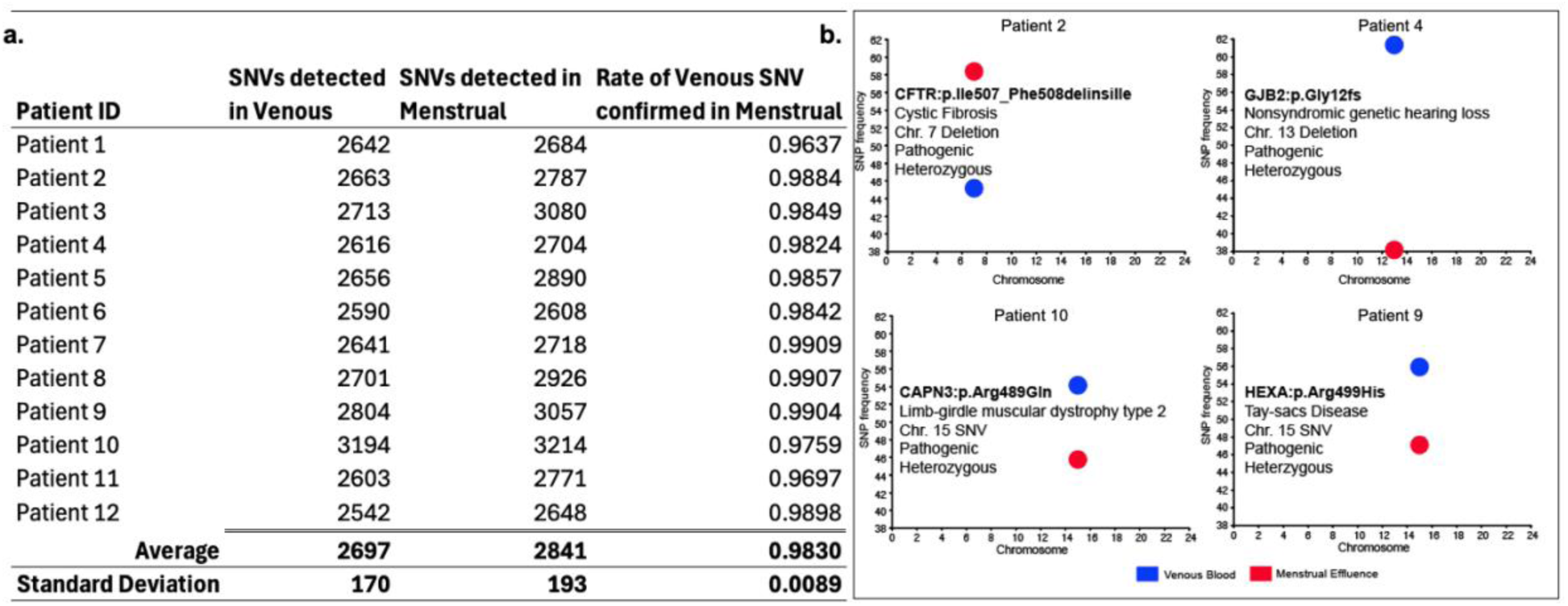
SNV detection in DNA from matched whole blood and menstrual effluence samples. SNV summary statistics for the comparison of SNVs between 12 patients who were exome sequenced using DNA extracted from menstrual effluence and from a venous blood draw. **a.** Table summarizing the number of SNVs recorded in the venous blood and menstrual effluence, and the rate of venous blood SNVs that were confirmed also in menstrual effluence. **b.** Four patients with known pathogenic mutations in CFTR, GJB2, CAPN3 and HEXA were confirmed present in both menstrual effluence and venous blood. Charts show mutation plotted against the frequency of the SNV detected. Red points are menstrual and blue points are venous. 24 samples from 12 participants were used in this analysis.

### The transcriptomic profile of menstrual effluence represents a unique composite of uterine, vaginal, and immune-derived cells distinct from isolated reproductive tissues

To assess tissue similarity, we compared RNA expression profiles of menstrual effluence collected via tampon (MB) and menstrual cup (MC) to GTEx data. Reproductive tissues (ovary, vagina, fallopian tubes, cervix, uterus), whole blood, and small intestine (as outgroup) were included. Spearman correlation was used to evaluate global similarity. Differential expression was performed between menstrual effluence and GTEx whole blood samples to determine overlapping genes between menstrual and reproductive tissues. The top 25 overexpressed genes in menstrual effluence show some overlap with reproductive tissues in GTEx (Figure 4b), demonstrating both shared and unique gene programs between the tissues. MB and MC samples were highly correlated (Spearman’s r = 0.97), with key differences in keratinized epithelium expression (Supplemental Figure 1a), likely due to vaginal wall cell inclusion in tampon samples^19^. Reproductive tissues showed low correlation (r = 0.23–0.32), comparable to small intestine or whole blood (r = 0.45; Figure 4a). This suggests that menstrual effluence shares only a limited transcriptional overlap with reproductive tissue biopsies, similar to the non-reproductive tissues included in the analysis. Differences in cycle phase and sequencing technology of GTEx data may also contribute to this low correlation. The mixture of blood, uterine, and vaginal tissue could represent shared gene programs between multiple tissue types leading to low correlations to any one tissue.

**Figure 4:**
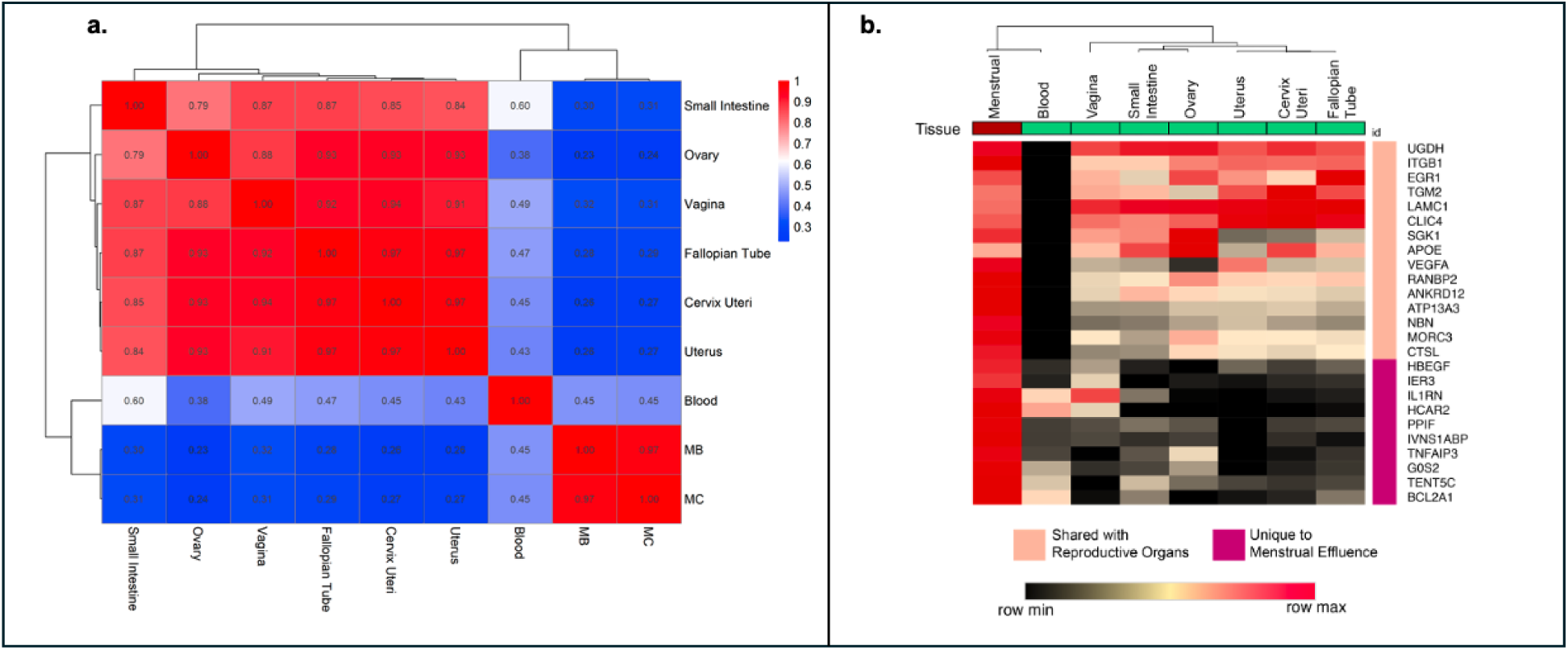
Sample type comparisons in menstrual effluence compared to other tissue types. Menstrual effluence gene expression global differences to other tissue types. **a.** Spearman correlation matrix of global RNA transcriptional activity in menstrual effluence (MB: tampon; MC: menstrual cup) compared to public datasets for small intestine, ovary, vagina, fallopian tube, cervix, uterus and blood. Red indicates a high level of transcriptional correlation between tissue types, while white and blue indicate a low level of correlation to other tissue types. 4,235 samples from GTeX and 218 samples from the NGJ dataset were used for this analysis. **b.** Top 25 differentially expressed genes between menstrual effluence and venous blood. Median gene values for 20 sequences from each GTeX tissue type were taken for comparison. Shared genes are grouped between reproductive tissue and menstrual (peach) vs menstrual samples with little to now expression in GTeX tissues (magenta). Expression differences in KRT13 between menstrual effluence collected in a menstrual cup or a tampon (p <0.0001; cohen’s f = 2.072). Analysis was done at the sequencing level (ESG) for 70 menstrual cup samples and 68 tampon samples.

### Gene expression signatures in menstrual effluence vary according to user-reported hormonal use, flow rate, and cycle timing, enabling molecular phenotyping

Endometrial gene expression provides valuable insight into menstrual physiology and molecular phenotypes, spanning objective metrics like flow rate (grams/hour) to subjective variables such as self-reported cycle day and flow type. To identify potential molecular correlates, we curated 89 endometrial-enriched genes from the Human Protein Atlas. Of these, 64 were detected in our sequencing data, with 30 genes showing JaneScore-normalized expression ≥2. Notable examples include canonical endometrial markers such as *PAEP*, *ESR1*, and several matrix metalloproteinases (*MMP10*, *MMP11*). The genes not present or at low expression may represent inherent differences between menstrual effluence and endometrium or be due to varying approaches in RNA sequencing.

Flow rate varies by birth control and subjective flow phenotype. Patients using any form of birth control exhibited reduced menstrual flow, as determined from 569 samples with reported data (Figure 5a). When flow rate was combined with spectrophotometric hemoglobin measurements at 550 nm, self-identified heavy bleeders (n = 83) showed significantly darker, more hemoglobin-rich effluence (p <0.0001; cohen’s f=0.352); Figure 5b). This single-tampon assay presents a scalable alternative to the burdensome alkaline hematin method, which requires collection of all products used over an entire cycle.

**Figure 5:**
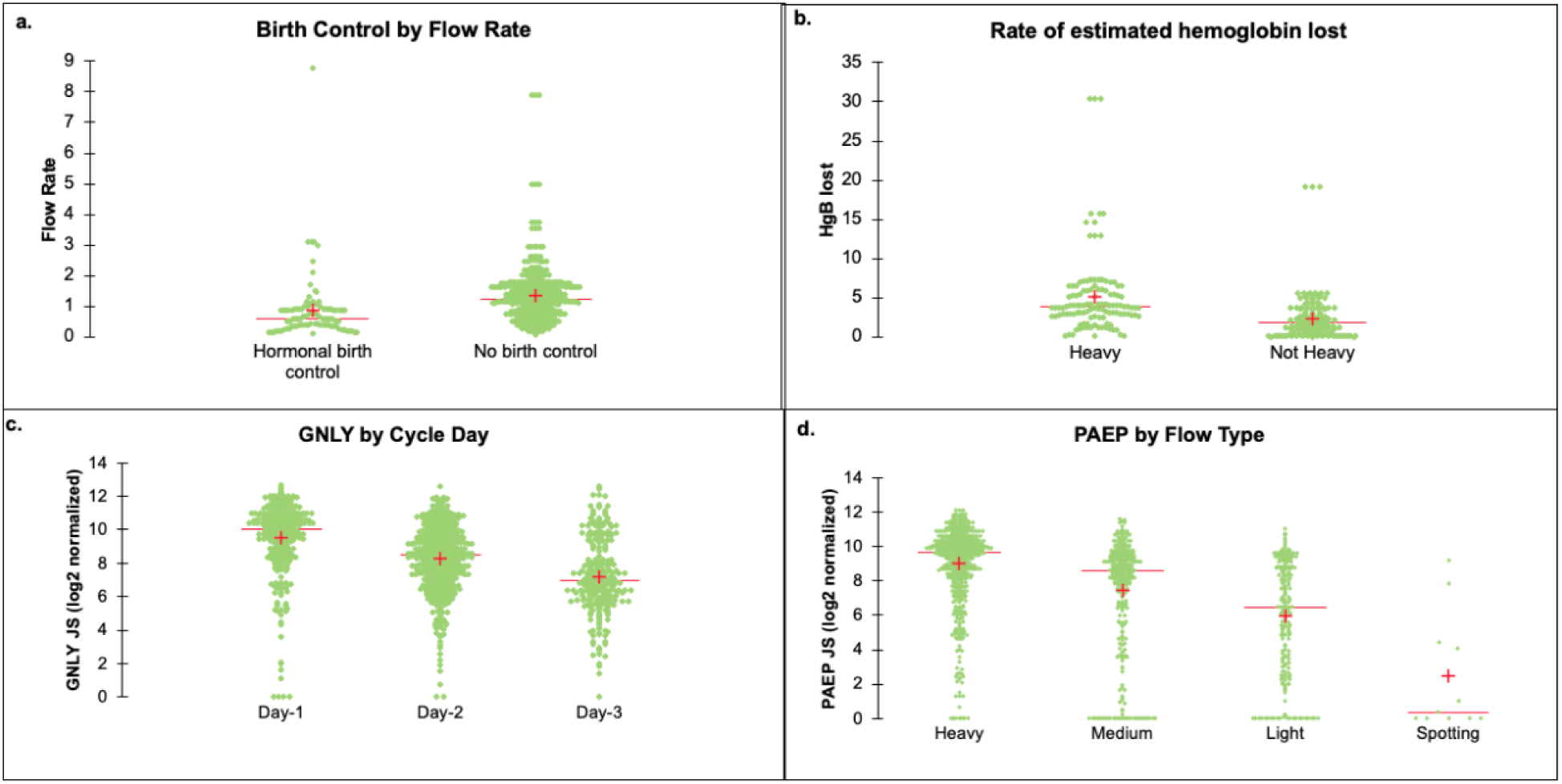
Flow rate and gene expression differences by sample collection characteristics. Flow rate and hemoglobin content by birth control and heavy menstrual bleeding and the differential expression of endometrial genes by cycle day. **a.** Flow rate calculated as grams of fluid collected per hour plotted against patient use of hormonal birth control. Patients on any form of hormonal birth control show lower flow rates (p <0.0001; cohen’s f = 0.204). Analysis was performed at the kit level for 241 tampons with recorded flow rate. **b.** Flow rate multiplied by the rfu (relative fluorescence units) spectrophotometer reading as a metric for hemoglobin within the sample charted by whether patients self-report as heavy menstrual bleeders or normal menstrual bleeders. Heavy bleeders show an increase in lost rate of hemoglobin per hour (p <0.0001; cohen’s f = 0.352). Analysis was performed at the kit level for 241 tampons with recorded flow rate. **c.** GNLY expression, an endometrium-enriched gene, by the day in the cycle (first cycle day represents first day of bleeding), shows significant expression changes by cycle day (p <0.0001; cohen’s f = 0.394). **d.** PAEP, another endometrium enriched gene charted by the day specific flow type during the tampon collection (p <0.0001; cohen’s f = 0.469). Mean and Median values are represented by a red “+” and “––”, respectively. C and d analyses were performed at the sequence level (ESG) for 1,135 samples.

Genes enriched in the endometrium correlate with cycle day and flow type. Expression of *GNLY*, a broad-spectrum antimicrobial protein, varied significantly by cycle day (p < 0.0001; cohen’s f=0.394; Figure 5c), indicating immune modulation across menstruation. Furthermore, self-reported flow heaviness (Heavy, Medium, Light, Spotting) was strongly associated with *PAEP* expression in tampon samples (p < 0.0001; cohen’s f= 0.469; Figure 5d).

### Menstrual effluence contains transcriptionally active microbial signatures that vary with menstrual day and may include clinically relevant pathogens

Menstrual samples show a time dependent dysbiosis in microbial transcription with an increase in BV causing bacteria on days 2 and 3 of menstruation. RNA transcriptional activity is distinct from the presence (relative abundance) of microbial DNA. We performed genus-level metatranscriptomic analysis using Kraken2 on RNA reads. We identified high transcriptional activity from several bacterial genera commonly associated with vaginal and uterine microbiota, including *Prevotella*, *Gardnerella*, *Corynebacterium*, *Finegoldia*, and *Anaerococcus* (Figure 6a). These genera are often implicated in microbial dysbiosis and reproductive tract infections. *Lactobacillus* species – typically associated with a healthy reproductive tract – were found to inversely correlate with the abundance of other bacterial taxa. In samples where *Lactobacillus* activity was high, the activity of other bacteria was generally suppressed. However, many samples exhibited low *Lactobacillus* levels (sum of all *Lactobacillus* species below 50% in 746 out of 1,135 sequences). alongside elevated expression of potentially pathogenic or opportunistic species. Interestingly, total *Lactobacillus* transcriptional activity decreased significantly on menstrual days 2 and 3 (Figure 6b), suggesting a temporal shift in microbial composition during menstruation. These findings present an opportunity to explore and understand typical microbial transitions during a single menstrual cycle and map how vaginal microbiome returns to equilibrium after a menstrual disruption.

**Figure 6:**
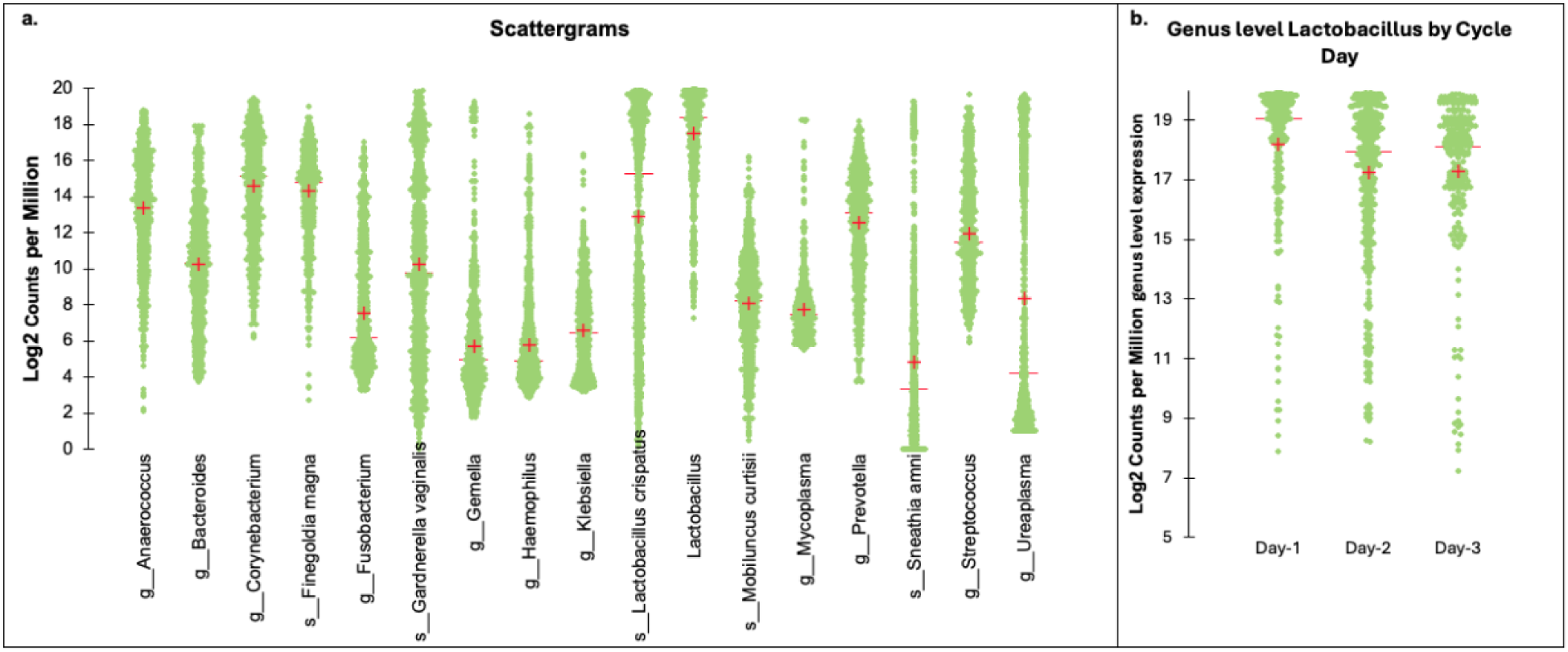
RNA transcriptional activity of key bacteria in menstrual effluence. **a.** *L. crispatus*, which is one beneficial lactobacillus species in the vaginal cavity alongside BV causing bacteria at the genus level. CPM counts per genus were summed before log2 transformation. Mean and Median values are represented by a “+” and “––”, respectively. **b.** Genus level *Lactobacillus* shows a significant decrease in transcriptional activity on day 2 and day 3 of menstruation, compared to day 1 (p <0.0001, cohen’s f = 0.171). Analyses were performed at the sequence level (ESG) for 1,135 samples.

## DISCUSSION

This study presents a standardized, at-home tampon-based system for the collection and stabilization of menstrual effluence, offering a scalable approach for molecular diagnostics and research in women’s health. The system preserves RNA and DNA for at least two weeks without refrigeration, supporting applications such as genetic carrier screening, RNA and exome sequencing, and microbial profiling. Its simplicity and suitability for at-home sampling help overcome persistent barriers to diagnostic access, especially in underserved populations. Through the NGJ platform, patients seeking to get pregnant could access their genetic screening results and uterine microbiome insight from a single sample collected at home.

A key contribution of this work is the ability to maintain nucleic acid integrity in a biologically complex, degradation-prone matrix. Menstrual effluence contains a heterogeneous mixture of blood, endometrial tissue, vaginal secretions, and immune cells, which contribute to enzymatic degradation. By limiting tampon wear to four hours, using chemically inert, low-absorbency cotton tampons, and stabilizing samples in a high-performance buffer, we preserved high-quality RNA and DNA for downstream molecular applications. Higher menstrual flow rates correlated with improved RNA quality, likely due to greater blood content and reduced degradation time.

Post-collection experiments further underscored the importance of immediate stabilization, as RNA integrity declined significantly in samples exposed to 37°C for just 30 minutes before preservation (Supplementary Figure 1). These findings provide actionable insights for real-world implementation of mail-in collection systems.

From a systems biology perspective, our findings highlight menstruation as a dynamic and immunologically active window into reproductive health. Transcriptomic profiles varied across the cycle and by hormonal use, with genes such as *PAEP*, *ESR1*, *GNLY*, and *MMP10* reflecting hormonal withdrawal, immune activation, and tissue remodeling. These changes are difficult to capture via biopsy or swabs, underscoring the value of menstrual effluence for temporal molecular profiling that should be accounted for in research and clinical findings^30–32^.

The high concordance between venous blood and menstrual effluence in exome sequencing supports its use as a non-invasive alternative for carrier screening. Variants linked to conditions such as cystic fibrosis and Tay-Sachs were concordantly detected in both sample types, demonstrating that menstrual effluence can expand access to genomic testing.

Our system also links subjective menstrual experiences with objective molecular measurements. Variables such as flow heaviness and cycle day correlated with RNA gene expression patterns, suggesting tampon-derived metrics could serve as proxies for subjective reporting or hematin-based methods in heavy menstrual bleeding classification.

Beyond transcriptomics, metatranscriptomic profiling revealed day-specific shifts in microbial communities. Decreases in *Lactobacillus* and increases in *Gardnerella* and *Prevotella* during menstruation suggest a transient dysbiosis. Coupling host transcriptional responses with microbial activity enables nuanced assessment of conditions like bacterial vaginosis or chronic endometritis, which affect a significant proportion of infertile individuals^33^. Immune activation markers may help determine whether antibiotic treatment is warranted in asymptomatic cases^34,35^. Our system allows for this dual measurement in a single, non-invasive specimen. RNA transcriptional profiles of the menstrual microbial community expand the potential of menstrual effluence as a non-invasive monitoring tool for chronic infections, sexually transmitted infections, or dysbiosis-associated conditions like pelvic inflammatory disease.

The reproducibility and standardization of this collection method also make it well-suited for research at scale. By capturing molecular profiles tied to menstrual phenotypes, this platform can enable the development of classifiers for endometriosis, fibroids, and other gynecologic conditions. Future work will endeavor to utilize this system to better understand the molecular characteristics of menstrual effluence and how variation in cycle data within patients and between patients may influence normal or abnormal menstruation.

## CONCLUSION

In conclusion, the development and validation of a standardized tampon-based system for menstrual effluence collection marks a significant advancement in women’s health diagnostics. Our findings demonstrate that menstrual effluence, a sample previously considered suboptimal for molecular analysis, can be preserved effectively for RNA and DNA extraction, enabling genetic screening, molecular phenotyping, and pathogen surveillance. This system offers unique opportunities for at-home, longitudinal monitoring of reproductive health, facilitating both clinical care and research applications.

The ability to link menstrual biomarkers with clinical outcomes, such as genetic carrier status or microbial dysbiosis, provides new avenues for early detection and personalized treatment strategies. By providing reliable, reproducible molecular data across diverse menstrual phases and bleeding phenotypes, this approach enhances the potential for diagnostic tools that address the unique health needs of women. Future work will focus on refining the system’s integration into clinical practice, developing molecular classifiers for reproductive diseases like endometriosis, and expanding the applications of menstrual effluence in broader health research.

## Supporting information

DNA seq metadata for SNV analysis

GTex Analysis sample list and Spearman table

RNA seq metadata and gene expression data

## Data Availability

All data produced in the present study are available upon reasonable request to the authors

## Acknowledgements

We would like to thank Stephen Palmer, Ruth Kulicke, Matt Stremlau, Ryan Phan, and Pardis Sabeti for their helpful edits. We would like to thank the LabCorp team that helped coordinate exome sequencing efforts: Marcia Eisenberg, Christina Smith, and Megann Vaughn. We would like to thank Abhishek Jha and Yogesh Lakhotia for bioinformatic support. Margaret Eisen, Julia Carr and Aparna Kola coordinated all clinical collection efforts for this study. The analyses in this paper were partly funded through an SBIR Fast Track grant with the National Institute of Child Health and Human Development *5R44HD103159*.

---

**Supplementary Table 1:** Sample filtering parameters. Menstrual samples collected on cycle days 1-3 that had complete metadata for collection date, bleeding phenotype, flow type, birth control, collection time and reproductive stage were included in the analysis. Samples were further filtered by technical sequencing QC metrics including proper strandedness of reads, ribosomal rRNA contamination and uniquely mapped reads.

|  |  |
| --- | --- |
| **All Samples** | **1867** |
| **Cycle day and Menstrual** | **1600** |
| Cycle days 1-3 |  |
| Exclude vaginal only samples |  |
| **Missing Data (0, Not Recorded, or NA)** | **1346** |
| Collection Date |  |
| Bleeding Phenotype |  |
| Flow type |  |
| Birth Control |  |
| Collection time |  |
| Reproductive Stage |  |
| **Technical Filters** | **1135** |
| Strandedness >0.8 |  |
| rRNA < 3% |  |
| Uniquely mapped reads >8M |  |

**Supplemental Figure 1:**
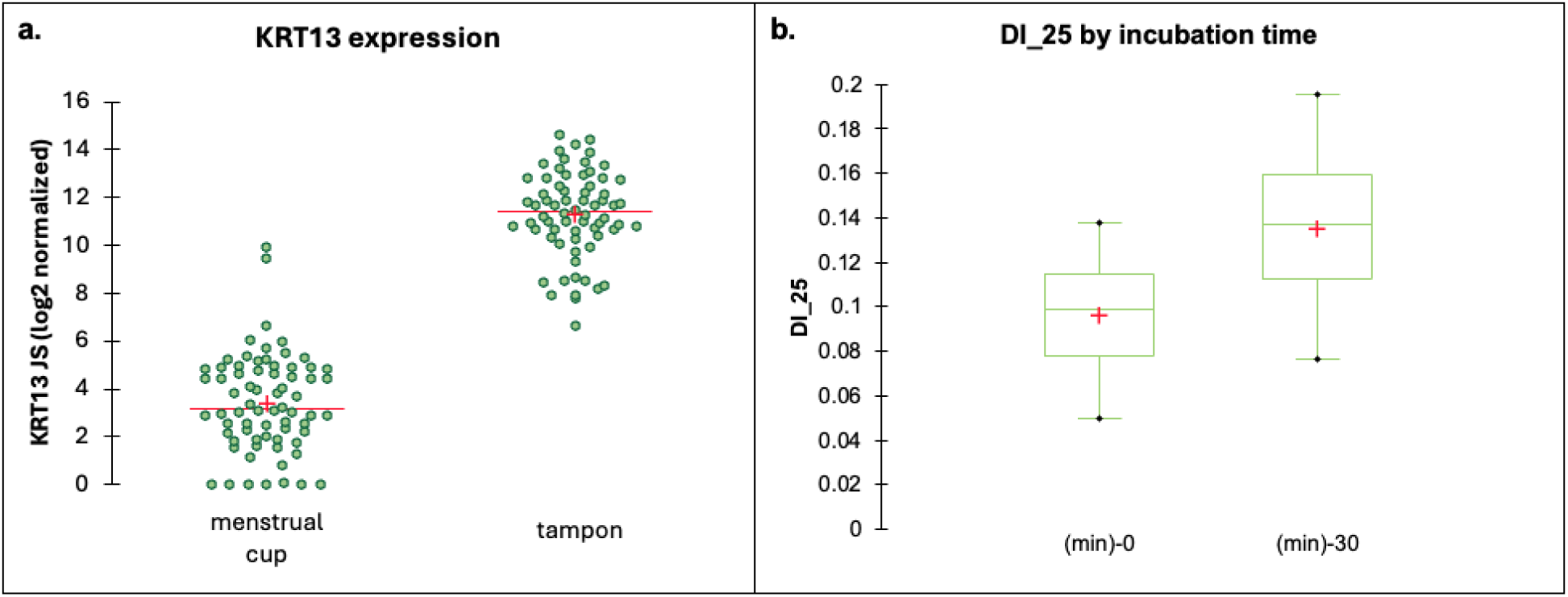
Supplemental analyses on menstrual cups. **a.** Expression differences in KRT13 between menstrual effluence collected in a menstrual cup or a tampon (p <0.0001; cohen’s f = 2.072). Analysis was done at the sequencing level (ESG) for 70 menstrual cup samples and 68 tampon samples. **b.** Menstrual effluence collected in menstrual cups was either directly put into preservation buffer after collection or incubated at 37oC for 30 minutes before preservation (p=0.003; cohen’s f = 0.606). A total of 10 diva cups from 5 participants were used in this analysis.

